# Communicating the benefits of quitting smoking on mental health increases motivation to quit in people with anxiety and/or depression: A randomized trial of two health messages

**DOI:** 10.1101/2023.02.17.23286103

**Authors:** Marc L. Steinberg, Rachel L. Rosen, Ollie Ganz, Olivia A. Wackowski, Michelle Jeong, Cristine D. Delnevo

## Abstract

**Background:** Although smoking rates have declined over time, this decline has not been observed among those with mental health concerns. It is therefore important to develop effective messaging to support quitting in this population.

**Methods:** We conducted an online experiment with 419 adults who smoke cigarettes daily. Participants with, or without a lifetime history of anxiety and/or depression were randomized to view a message focused on the benefits of quitting smoking on mental or physical health. Participants then reported motivation to quit smoking, mental health concerns about quitting, and perceived effectiveness of the message.

**Results:** Participants with a lifetime history of anxiety and/or depression who saw the message focused on the benefits of quitting smoking on mental health reported greater motivation to quit than when they saw a message focused on the benefits to physical health. This was not replicated when examining current symptoms instead of lifetime history. Pre-existing beliefs that smoking improves one’s mood were greater in those experiencing current symptoms and in those with a lifetime history of anxiety and/or depression. There was no main or interaction (message type X mental health status) effect of message type received on mental health related concerns about quitting.

**Conclusions:** This study is one of the first to evaluate a smoking cessation message with content specifically targeted to those with mental health concerns about quitting smoking. Additional work is needed to determine how to best target those with mental health concerns with messages focused on the benefits of quitting on mental health.

**Implications:** These data can begin to inform regulatory efforts to address tobacco use in those with comorbid anxiety and/or depression by providing information on how to effectively communicate the benefits of quitting smoking on mental health.

## INTRODUCTION

Although smoking rates have declined over time in the general population, this decline has not been observed in those with mental health concerns.[1,2] Relatedly, between 2008 and 2016 in the US, people who smoke cigarettes and report serious psychological distress, were approximately half as likely to quit smoking as those without serious psychological distress.[3] This contributes to significantly higher rates of cigarette smoking among adults with mental health concerns as compared to those without.[1,2,4] Data from the National Epidemiologic Survey on Alcohol and Related Conditions-III [5] reveal that after controlling for sociodemographic characteristics, those with affective disorders were more likely to currently smoke cigarettes than adults without mental health concerns.[6]

Higher rates of smoking among those with anxiety and/or depression may be related to beliefs that cigarette smoking can influence mood. Specifically, people who smoke cigarettes generally believe that smoking can reduce negative moods,[7,8] and one’s beliefs about the mood-enhancing effects of smoking not only influence decisions about making quit attempts,[9] but also predict successful quitting.[10] This is relevant to individuals with current symptoms and to those with a history of anxiety and/or depression as it is not uncommon for these disorders to recur.[11] If individuals who smoke cigarettes believe that quitting will make their mental health suffer (including precipitating a recurrence of past symptoms), they may be less likely to make a quit attempt. Indeed, many clinicians and people who smoke cigarettes believe that smoking is an important coping mechanism for those with mental health concerns.[12] Notably, the tobacco industry has perpetuated this belief by promoting psychological benefits of smoking in their advertising.[13]

The literature, however, does not support this view that quitting smoking will exacerbate mental health symptoms. A systematic review and meta-analysis of 26 longitudinal studies found that symptoms of anxiety, depression, mixed anxiety and depression, and stress significantly decreased in those who quit smoking compared to those who did not.[14] Similarly, psychological quality of life and positive affect significantly increased in those who quit compared to those who did not.[14]

One approach for correcting this misperception regarding the effects of quitting smoking and mental health is to create public health campaigns or to provide individual corrective messaging. Although the U.S. Food and Drug Administration (FDA) has created anti-tobacco public health campaigns with messages targeted for teens,[15] at-risk youth identifying with hip-hop culture,[16] lesbian, gay, bisexual and transgender young adults,[17] and those who have made unsuccessful quit attempts,[18] no FDA public education campaigns have been targeted to adults with mental health concerns (a yet to be studied campaign targeted towards the negative mental health effects of smoking on teens was released in January 2023[19]). One federal initiative, the Centers for Disease Control and Prevention’s (CDC) Tips From Former Smokers (Tips) campaign, [20] included a message targeting adults with mental health concerns as part of a larger campaign; however, there are currently no federal campaigns designed to communicate the message that, consistent with research,[14] individuals with mental health concerns who smoke can quit smoking without risk of exacerbating their psychiatric symptoms and, further, that mental health symptoms may actually improve with abstinence.

A systematic review [21] examining the influence of anti-smoking public health messages on those with mental health concerns identified only eight studies. The only study in the review that examined a public health message that included a specific reference to mental health (i.e., “Rebecca’s Tip” from the CDC’s Tips ad campaign [22,23]) highlighted an important and overlooked effect of message exposure. Specifically, a greater frequency of exposure to “Rebecca’s Tip”, was associated with greater odds of making a quit attempt during the campaign and greater intention to quit smoking cigarettes for those with, but not without, a history of self-reported mental health disorders.[24] In contrast, greater frequency of exposure to ads that primarily referenced physical health benefits of quitting smoking was associated with greater odds of making a quit attempt among those without, but not with, a history of mental health disorders.[24]

Likewise, a recent review of tobacco regulatory science research [25] reported a gap in the literature regarding how health communication influences tobacco use among those with mental health concerns. Given that the proportion of people who smoke cigarettes who have mental health concerns is increasing over time,[1–3] it is especially important to develop effective messaging to support quitting in this population. Existing messaging focused primarily on physical health effects of smoking, which are common in most large-scale, public education campaigns, appear to be ineffective for people with mental health concerns and there is emerging evidence that messages targeted to those with mental health concerns may be effective. To fill this research gap, we examined beliefs about the effects of tobacco on mental health and how messaging related to these beliefs influences motivation to quit smoking and concerns about quitting smoking among those with a lifetime history of anxiety and/or depression. We hypothesized that 1) People who smoke cigarettes with, as compared to without a lifetime history of anxiety and/or depression will more strongly endorse beliefs regarding cigarette smoking as a mood enhancer and 2) People who smoke cigarettes with a lifetime history of anxiety and/or depression (vs. those without) who view a message related to improved psychological (vs. physical) effects of quitting smoking will report increased motivation to quit and lower ratings of concerns about the impact of quitting on mental health as compared to all other groups.

## METHODS

### Sample and Experimental Design

Participants were recruited by the research company Ipsos from their KnowledgePanel®, the largest online panel in the US that relies on probability-based sampling methods for recruitment to provide a representative sampling frame for adults in the US. KnowledgePanel® uses an address-based sampling methodology, which is independent of respondents’ phone status. Individuals were screened for a lifetime history of anxiety and/or depression to recruit approximately half the sample with and half without a lifetime history of anxiety and/or depression. Approximately half the sample (n = 211) reported being told by a doctor or other health professional that they have any type of depression or an anxiety disorder and approximately half (n = 208) reported that they have NOT been told this. Furthermore, inclusion criteria included being at least 18 years old, smoking cigarettes daily, residing in the US, and being able to read English.

After providing informed consent, the full sample of participants (N = 419) were randomly assigned in a 1:1 ratio to receive a health message related to either the physical or the mental health benefits of quitting smoking. All participants completed measures of cigarette dependence, physical and mental health, cigarette smoking beliefs, and demographic charactieristics before viewing the message to which they were randomly assigned. After viewing the message, participants answered items regarding motivation to quit smoking cigarettes, concerns about quitting smoking, and perceived effectiveness of the messages. Ipsos administered the survey and online experiment in March 2022, and the study was approved by the Rutgers Health Sciences IRB.

The health messages (see Supplemental File 1) were developed for this study, and each included 128 words and were presented on a single survey page. They were rated as 8.76 (physical health) and 8.88 (mental health) on the Gunning Fog Index [26] for readability indicating that the messages were written at an approximately 8^th^ grade reading level. The index score was calculated via Online-Utility.org.[27]

### Measures

Prior to message exposure, participants provided information on demographic characteristics, cigarette dependence, physical and mental health, and beliefs about cigarette smoking as described below.

#### Cigarette Dependence

The Heaviness of Smoking Index (HSI [28,29]) is a commonly used measure of cigarette dependence based on number of cigarettes smoked per day and the time-to-first-cigarette of the morning.

#### Physical and Mental Health

In addition to being screened for the presence or absence of a mental health history as part of the study’s inclusion criteria, participants also reported current severity of depression with the Patient Health Questionnaire (PHQ-9 [30]) and severity of anxiety with the Generalized Anxiety Disorder-7 (GAD-7 [31]). The PHQ-9 and GAD-7 items are rated on a scale of 0 to 3 and total scores are computed by summing their items with possible scores ranging from a low of 0 to a high of 27 (for the PHQ-9) or 21 (for the GAD-7). We provided all participants with contact information for the National Suicide Prevention Lifeline because the PHQ-9 assessed for suicidal ideation. Finally, we included a single item assessing general physical health (“*Would you say your health in general is excellent, very good, good, fair, or poor?*”).

#### Cigarette smoking beliefs

We assessed participants’ beliefs about the mood enhancing effects of cigarette smoking via items from the Wisconsin Inventory of Smoking Dependence Motives (WISDM-68 [32,33]), a widely used measure with strong psychometric properties designed to elucidate smoking motives that contribute to dependence. The items were chosen a priori as relevant to beliefs that smoking cigarettes could improve one’s mood. The items included all six from the original WISDM’s [32] Negative Reinforcement scale and one from the Positive Reinforcement scale. Three of these items represented the full Affective Enhancement Scale of the Brief WISDM.[33] These items were completed before participants viewed the health messages.

After exposure to the health message, participants provided information about motivation to quit smoking, concerns about quitting smoking, and their perceived effectiveness of the messages as described below.

#### Motivation to Change

Participants described their motivation to quit on the 3-item Change Measure [34] in which participants responded to the items, “It is important for me to quit smoking”, “I am trying to quit smoking”, and “I could quit smoking” using a 10-point Likert scale. We calculated a total motivation score by summing scores across the three items.

#### Concerns About Quitting

Participants responded to six questions about concerns of quitting related to one’s mental health. We generated a total score by calculating the average across all six items (e.g., “If I quit smoking I will experience [sadness, anxiety, stress, problems sleeping, mental health symptom worsening, irritability].”

#### Perceived Effectiveness

Participants rated the messages’ effectiveness using a 6-item scale that has commonly been used in pre-testing FDA campaigns.[35–37] We generated a total Perceived Effectiveness score by calculating the average across the six items.

### Analytic Plan

Data were weighted to be nationally representative and to adjust for sampling. Throughout the text, we present unweighted sample sizes and weighted population estimates. To examine beliefs relating to cigarette smoking as a mood enhancer in people with and without anxiety and/or depression, we conducted a linear regression analysis with diagnosis as the independent variable and average rating of selected WISDM-68 items (i.e., beliefs about smoking) as the dependent variable. We included nicotine dependence as measured by the HSI score as a covariate. To determine the effects of physical and psychological health messages on 1) perceived harms to mental health from quitting, 2) perceived message effectiveness, and 3) motivation to stop cigarette use in people with and without anxiety and/or depression, we conducted separate linear regression analyses with history of depression and/or anxiety diagnosis and message type (and their interaction) as independent variables. Perceived risk to mental health from quitting smoking, perceived message effectiveness, and motivation to quit were included as dependent variables. We included the HSI as a covariate in the analysis of motivation to quit. Because lifetime history of anxiety and/or depression does not capture current impairment/distress associated with anxiety and depression and may be influenced by other factors, such as access to mental health care, we conducted sensitivity analyses to examine the effect of current symptoms, assessed via the GAD-7 and PHQ-9, on all primary outcomes. Anxiety (GAD-7) and depression (PHQ-9) symptom scores were highly correlated (*r* = 0.85, *p* < 0.001) so we examined their effects in separate sets of analyses. Table 1 displays participant characteristics and all weighted regression models are displayed in Table 2.

**Table 1.**
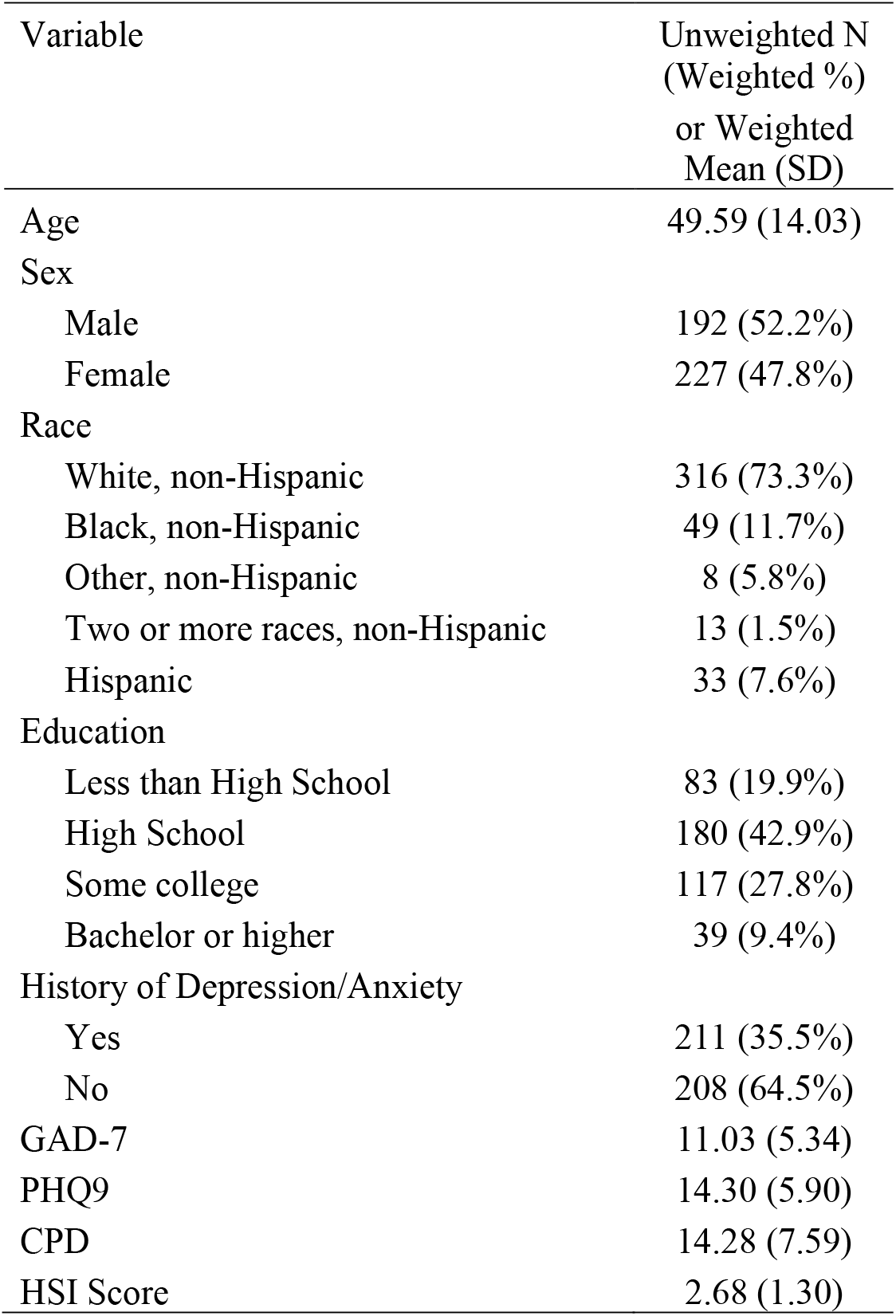
Participant characteristics (N=419)

| Variable | Unweighted N<br>(Weighted %)<br>or Weighted<br>Mean (SD) |
| --- | --- |
| Age | 49.59 (14.03) |
| Sex |  |
| Male | 192 (52.2%) |
| Female | 227 (47.8%) |
| Race |  |
| White, non-Hispanic | 316 (73.3%) |
| Black, non-Hispanic | 49 (11.7%) |
| Other, non-Hispanic | 8 (5.8%) |
| Two or more races, non-Hispanic | 13 (1.5%) |
| Hispanic | 33 (7.6%) |
| Education |  |
| Less than High School | 83 (19.9%) |
| High School | 180 (42.9%) |
| Some college | 117 (27.8%) |
| Bachelor or higher | 39 (9.4%) |
| History of Depression/Anxiety |  |
| Yes | 211 (35.5%) |
| No | 208 (64.5%) |
| GAD-7 | 11.03 (5.34) |
| PHQ9 | 14.30 (5.90) |
| CPD | 14.28 (7.59) |
| HSI Score | 2.68 (1.30) |

**Table 2.**
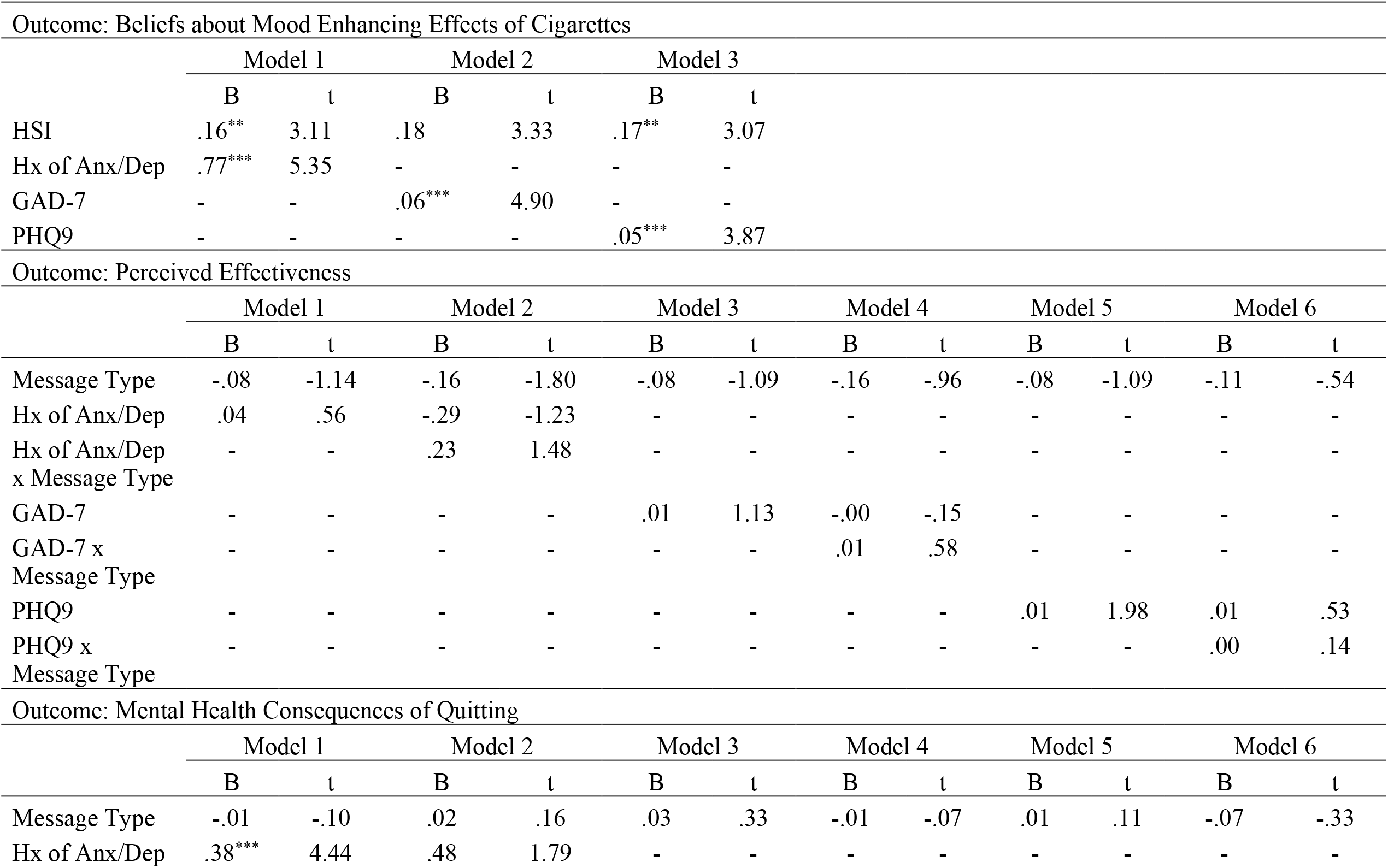
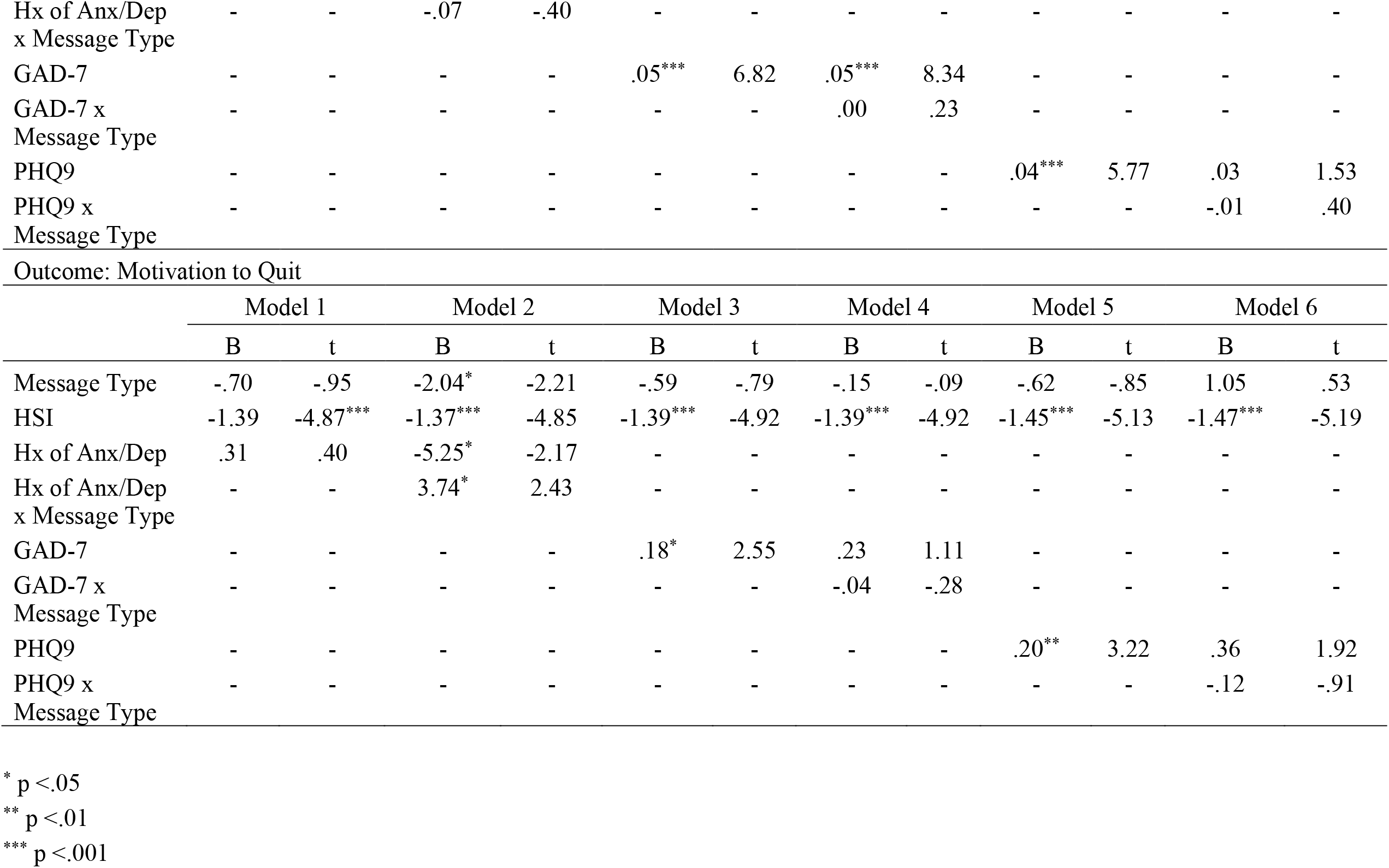
Weighted regression models.

| Outcome: Beliefs about Mood Enhancing Effects of Cigarettes |  |  |  |  |  |  |  |  |  |  |  |  |
| --- | --- | --- | --- | --- | --- | --- | --- | --- | --- | --- | --- | --- |
|  | Model 1 |  | Model 2 |  | Model 3 |  |  |  |  |  |  |  |
|  | B | t | B | t | B | t |  |  |  |  |  |  |
| HSI | .16** | 3.11 | .18 | 3.33 | .17** | 3.07 |  |  |  |  |  |  |
| Hx of Anx/Dep | .77*** | 5.35 | - | - | - | - |  |  |  |  |  |  |
| GAD-7 | - | - | .06*** | 4.90 | - | - |  |  |  |  |  |  |
| PHQ9 | - | - | - | - | .05*** | 3.87 |  |  |  |  |  |  |
| Outcome: Perceived Effectiveness |  |  |  |  |  |  |  |  |  |  |  |  |
|  | Model 1 |  | Model 2 |  | Model 3 |  | Model 4 |  | Model 5 |  | Model 6 |  |
|  | B | t | B | t | B | t | B | t | B | t | B | t |
| Message Type | -.08 | -1.14 | -.16 | -1.80 | -.08 | -1.09 | -.16 | -.96 | -.08 | -1.09 | -.11 | -.54 |
| Hx of Anx/Dep | .04 | .56 | -.29 | -1.23 | - | - | - | - | - | - | - | - |
| Hx of Anx/Dep<br>x Message Type | - | - | .23 | 1.48 | - | - | - | - | - | - | - | - |
| GAD-7 | - | - | - | - | .01 | 1.13 | -.00 | -.15 | - | - | - | - |
| GAD-7 x<br>Message Type | - | - | - | - | - | - | .01 | .58 | - | - | - | - |
| PHQ9 | - | - | - | - | - | - | - | - | .01 | 1.98 | .01 | .53 |
| PHQ9 x<br>Message Type | - | - | - | - | - | - | - | - | - | - | .00 | .14 |
| Outcome: Mental Health Consequences of Quitting |  |  |  |  |  |  |  |  |  |  |  |  |
|  | Model 1 |  | Model 2 |  | Model 3 |  | Model 4 |  | Model 5 |  | Model 6 |  |
|  | B | t | B | t | B | t | B | t | B | t | B | t |
| Message Type | -.01 | -.10 | .02 | .16 | .03 | .33 | -.01 | -.07 | .01 | .11 | -.07 | -.33 |
| Hx of Anx/Dep | .38*** | 4.44 | .48 | 1.79 | - | - | - | - | - | - | - | - |
| RUNNING HEAD: MESSAGING ABOUT TOBACCO AND MENTAL HEALTH |  |  |  |  |  |  |  |  |  |  |  |  |
| Hx of Anx/Dep<br>x Message Type | - | - | -.07 | -.40 | - | - | - | - | - | - | - | - |
| GAD-7 | - | - | - | - | .05*** | 6.82 | .05*** | 8.34 | - | - | - | - |
| GAD-7 x<br>Message Type | - | - | - | - | - | - | .00 | .23 | - | - | - | - |
| PHQ9 | - | - | - | - | - | - | - | - | .04*** | 5.77 | .03 | 1.53 |
| PHQ9 x<br>Message Type | - | - | - | - | - | - | - | - | - | - | -.01 | .40 |
| Outcome: Motivation to Quit |  |  |  |  |  |  |  |  |  |  |  |  |
|  | Model 1 |  | Model 2 |  | Model 3 |  | Model 4 |  | Model 5 |  | Model 6 |  |
|  | B | t | B | t | B | t | B | t | B | t | B | t |
| Message Type | -.70 | -.95 | -2.04* | -2.21 | -.59 | -.79 | -.15 | -.09 | -.62 | -.85 | 1.05 | .53 |
| HSI | -1.39 | -4.87*** | -1.37*** | -4.85 | -1.39*** | -4.92 | -1.39*** | -4.92 | -1.45*** | -5.13 | -1.47*** | -5.19 |
| Hx of Anx/Dep | .31 | .40 | -5.25* | -2.17 | - | - | - | - | - | - | - | - |
| Hx of Anx/Dep<br>x Message Type | - | - | 3.74* | 2.43 | - | - | - | - | - | - | - | - |
| GAD-7 | - | - | - | - | .18* | 2.55 | .23 | 1.11 | - | - | - | - |
| GAD-7 x<br>Message Type | - | - | - | - | - | - | -.04 | -.28 | - | - | - | - |
| PHQ9 | - | - | - | - | - | - | - | - | .20** | 3.22 | .36 | 1.92 |
| PHQ9 x<br>Message Type | - | - | - | - | - | - | - | - | - | - | -.12 | -.91 |

\* p < .05
\*\* p < .01
\*\*\* p < .001

## RESULTS

### Descriptive Statistics

On average, individuals were 49.59 years old (SD=14.03). About half were male (N=192, 52.2%). Most individuals identified as White, non-Hispanic (N=316, 73.3%), while 11.7% (N=49) were Black, non-Hispanic. Approximately one-third (N=184, 31.4%) had been diagnosed with a depressive disorder and one-quarter (N=162, 27.3%) had been diagnosed with an anxiety disorder in their lifetime. One-quarter (N=135, 23.1%) had a history of both anxiety and depression. On average, individuals reported moderate levels of depression (M=14.30, SD=5.90) and anxiety (M=11.02, SD=5.34) based on the PHQ-9 and GAD-7, respectively. Individuals with a lifetime history of anxiety and/or depression had significantly higher current symptoms of anxiety (M=14.55, SD=6.11) and depression (M=18.03, SD=6.71) than those without a lifetime history of anxiety and/or depression (anxiety: M=9.07, SD=3.61; depression: M=12.23, SD=4.16, all ps< 0.001). Individuals smoked an average of 14.28 cigarettes per day (SD=7.59).

### Pre-message beliefs regarding cigarette smoking as a mood enhancer

A linear regression analysis showed that the effect of a lifetime history of anxiety and/or depression diagnosis was significantly associated with WISDM scores [*F*(2, 415)=20.19, *p*<.001)], controlling for HSI score; a history of anxiety and/or depression was associated with a .77 unit increase in WISDM score (*t*=5.35, *p*<.001). Separate analyses examining the effect of current symptoms of anxiety [*F*(2, 413)=17.79, *p*<.001)] and depression [*F*(2, 413)=13.15, *p*<.001)] on WIDSM scores, controlling for HSI were consistent with the above findings. Every one unit increase on the GAD-7 was associated with an .06 unit increase in WISDM score (*t*=4.90, *p*<.001) and every one unit increase on the PHQ9 was associated with a .05 unit increase in WISDM score (*t*=3.87, *p*<.001).

### Messaging

Separate linear regression analyses were conducted to examine the effect of lifetime history of an anxiety and/or depression diagnosis and tobacco cessation messaging (and their interaction) on 1) perceived effectiveness of the message, 2) Mental health concerns about quitting smoking, and 3) motivation to quit smoking (controlling for HSI score).

### Perceived effectiveness of messages

The mental health message’s perceived effectiveness was rated as 3.46 (SD=0.74) and the physical health message’s perceived effectiveness was rated as 3.58 (SD=0.75). We did not observe a significant effect lifetime history of anxiety and/or depression (nor current anxiety or depression symptoms), message type, or their interaction on perceived message effectiveness (all *p*>0.05).

### Post-message mental health concerns about quitting smoking

There was a significant effect of lifetime history of anxiety and/or depression on concerns about quitting smoking [*F*(2, 414)=9.84, *p*<.001)]. A history of anxiety and/or depression was associated with a .376 unit increase on the concerns about quitting measure (*t*=4.44, *p*<.001). There was no significant effect of message type received and no significant interaction effect (all *p*>.05). Separate analyses examining the effect of current symptoms of anxiety [*F*(2, 413)=23.22, *p*<.001)] and depression [*F*(2, 413)=16.64, *p*<.001)] on concerns about quitting smoking, controlling for HSI were consistent with the above findings. Every one unit increase on the GAD-7 was associated with an .05 unit increase on the concerns about quitting measure (*t*=6.82, *p*<.001) and every one unit increase on the PHQ9 was associated with a .04 unit increase on the concerns about quitting measure (*t*=5.77, *p*<.001). As above, there was no significant effect of message type and no significant interaction effect (all *p*>.05).

### Post-message motivation to quit

Consistent with hypotheses, there was a significant interaction of message type and lifetime history of anxiety and/or depression on motivation to quit (*B*=3.74, *t*=2.23, *p*=.016). Individuals with a history of depression and/or anxiety who saw a mental health message (M=16.28, SD=7.53) had higher motivation to quit smoking than if they saw a physical health message (M=14.43, SD=7.89). The inverse was true for individuals without a history of anxiety and/or depression, such that individuals reported higher motivation to quit smoking if they viewed a physical (M=16.24, SD=7.98) versus a mental health message (M=14.19, SD=7.46). There was a significant effect of HSI score on motivation to quit (p<.001), but there was not a significant effect of lifetime history of anxiety and/or depression or message type on motivation to quit, controlling for HSI score.

Separate analyses examining the effect of current symptoms of anxiety [*F*(3, 412)=10.38, *p*<.001)] and depression [*F*(3, 412)=11.75, *p*<.001)] on motivation to quit, controlling for HSI, provided slightly different findings. Specifically, there was a significant effect of current symptom severity on motivation to quit. Every one unit increase on the GAD-7 was associated with an .18 unit increase in motivation to quit (*t*=2.55, *p*=.01) and every one unit increase on the PHQ9 was associated with a .20 unit increase in motivation to quit (*t*=3.22, *p*=.001). There was no significant interaction between current symptoms and message type on motivation to quit (i.e., anxiety*message type; depression*message type), all *p*>.05).

## DISCUSSION

This is one of the first studies to evaluate a smoking cessation message with content specifically targeted to those with mental health concerns. As hypothesized, we found that individuals who smoke daily and have a lifetime history of anxiety and/or depression who saw a message focused on the mental health benefits of quitting smoking reported greater motivation to quit smoking than when they saw a message focused on the physical benefits. Correspondingly, those without a lifetime history of anxiety and/or depression reported greater motivation to quit smoking when they viewed a message focused on the physical, rather than the mental health benefits. It should be noted that while these differences were statistically significant, it is unclear how clinically significant they are and how they may influence later behavior.

These findings are consistent with those resulting from one related examination of ads from the CDC’s Tips ad campaign [20] in which greater exposure to an ad featuring a woman with depression (the “Rebecca” ad) by individuals with mental health concerns was associated with greater intention to quit smoking and a greater chance of making a quit attempt. Those without mental health concerns, however, were more influenced by other ads.[24] Unlike the messages tested in this study, the “Rebecca” ad campaign did not address mental health-specific beliefs about smoking or concerns about quitting, but rather simply provided backstories of individuals who had successfully quit smoking. It is possible, however, that the “Rebecca” ad and the mental health message in the current study had their influence on intention or motivation to quit by making the idea of quitting smoking seem more feasible to those with mental health concerns. Additionally, reading about the mental health benefits of quitting smoking in the current study may have challenged pre-existing beliefs about the role of smoking on mood management (among those with histories of anxiety and/or depression).

Findings from our sensitivity analysis examining current symptom severity instead of lifetime history showed that the interaction effect between current symptoms (rather than lifetime history) and message type (mental vs. physical symptoms) on motivation to quit smoking was not statistically significant. As expected, we found more severe levels of current anxiety and/or depression symptoms among those with, as compared to without, a lifetime history of anxiety and/or depression diagnoses. There was a higher degree, however, of anxiety and depression symptom severity than expected. In fact, even those denying a lifetime history of anxiety reported mild/moderate current symptoms of anxiety and those denying a lifetime history of depression reported moderate levels of depression. Consequently, there may not have been sufficient variation in symptomatology to detect an interaction effect when running the analyses by current symptoms instead of lifetime history. Alternatively, it may simply be that lifetime history of anxiety and/or depression is more relevant to how one responds to health messages related to the mental health benefits of quitting than current symptoms. It is reasonable to assume that even someone with relatively low levels of current mental health symptoms, but with a positive lifetime history, could be concerned about a mental health relapse if they were to quit smoking. Messaging about the mental health benefits of quitting smoking could allay those concerns about a mental health relapse.

Pre-existing beliefs that smoking improves one’s mood were greater in those with a lifetime history of anxiety and/or depression and in those with more severe symptoms of anxiety and/or depression. This is consistent with previous findings that those with, compared to those without, major depressive disorder, scored higher on the WISDM Affective Enhancement scale.[38,39] Given that these beliefs are held by people who smoke who have mental health concerns, messaging that directly targets these beliefs may be indicated.

Similarly, we found that mental health related concerns about quitting smoking were greater in those with more severe symptoms of, and a lifetime history of, anxiety and/or depression. There was no effect of message type received on mental health related concerns about quitting and no significant interaction between message type and mental health (lifetime history or current symptoms) on mental health related concerns. The lack of interaction effect may be due, in part, to the fact that participant ratings of our mental health messages’ perceived effectiveness were on the lower end of those reported by adults who smoke rating other cessation-related messages.[40] The perceived effectiveness ratings of the individual physical health focused ads associated with the Tips campaign [20] ranged from 3.47 to 3.80,[40] while participants in the current study rated the mental health message’s perceived effectiveness as 3.46 on the same scale. It is possible that our mental health message needs to be improved. It is also possible, though less likely, that despite our assumption that any differences in motivation to quit would be driven by reduced mental health concerns about quitting that our message influenced motivation to quit via a different mechanism.

An additional possible explanation for the lack of a statistically significant interaction effect is that some items we considered mental health concerns about quitting (e.g., quitting smoking resulting in poor sleep or increased stress) are also likely to be concerns held by individuals who smoke who do not have a have current symptoms, or a history of mental health concerns. Additionally, participants were only exposed to the message once. Campaign evaluation studies show that message impact on tobacco-related attitudes and beliefs is associated with frequency of exposure.[41] Future studies should examine the impact of multiple exposures to messages on mental health related concerns about quitting.

Study limitations include the use of a single message in each condition type and single message exposure. Additionally, while the messages fairly closely, it is possible that aspects of the message presentation could have influenced the study outcomes. Our sample is representative of the US population; however, our findings may not be as relevant for other countries with different demographic compositions, different views of mental health and the relationship between cigarette smoking and mental health, or different rates of current symptoms.

## Conclusions

This study of adults who smoke suggests that mental health concerns are related to beliefs about the connection between cigarette smoking and mood. Furthermore, messages addressing the mental health benefits of quitting smoking may increase motivation to quit among those with a history of anxiety and/or depression. These data can begin to inform regulatory efforts that may help the FDA to address tobacco use in those with anxiety and/or depression by providing information on how to effectively communicate the mental health benefits of quitting smoking.

## Supporting information

Supplemental file - Health Messages

## Data Availability

All data produced in the present study are available upon reasonable request to the authors.

## Notes

Research reported in this publication was supported by NCI and FDA Center for Tobacco Products (CTP) under Award Number U54CA229973. The content is solely the responsibility of the authors and does not necessarily represent the official views of the NIH or the Food and Drug Administration.

### Competing Interest Statement

The authors have declared no competing interest.

### Author Declarations

The Rutgers Health Sciences IRB of Rutgers University gave ethical approval for this work.

