## Supplemental file - Health Messages for "Communicating the benefits of quitting smoking on mental health increases motivation to quit in people with anxiety and/or depression: A randomized trial of two health messages"

Quitting smoking is one of the most important steps people can take to improve their physical health. This is true no matter how old they are or how long they have been smoking![1]

#### Quitting smoking:[1]

**Benefit 1**

Increases life expectancy.

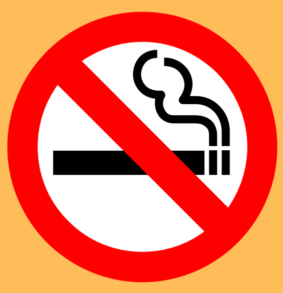

**Benefit 2**

Lowers the risk of 12 types of cancer.

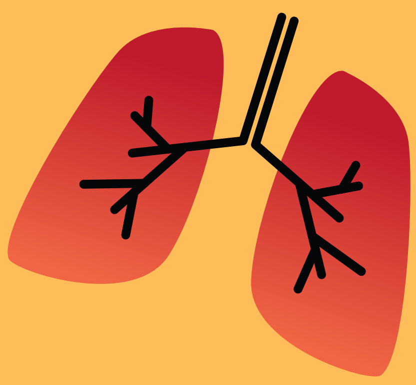

**Benefit 3**

Lowers the risk of heart disease.

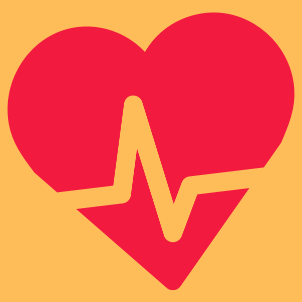

**Benefit 4**

Lowers the risk of COPD.

**Benefit 5**

Improves health status.

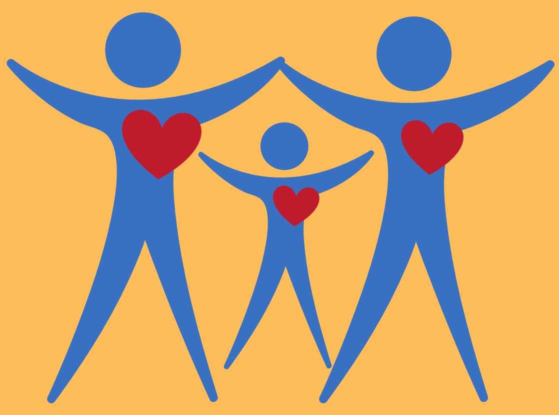

The good news is that it is never too late to quit smoking. Quitting smoking benefits physical health at any age. Even heavy smokers and people who have smoked for many years will benefit from quitting.[1]


1. U.S. Department of Health and Human Services. Smoking Cessation: A Report of the Surgeon General. Atlanta, GA: U.S. Department of Health and Human Services, Centers for Disease Control and Prevention, National Center for Chronic Disease Prevention and Health Promotion, Office on Smoking and Health, 2020.[accessed 2020 May 13].
2. U.S. Department of Health and Human Services. The Health Consequences of Smoking—50 Years of Progress: A Report of the Surgeon General. Atlanta: U.S. Department of Health and Human Services, Centers for Disease Control and Prevention, National Center for Chronic Disease Prevention and Health Promotion, Office on Smoking and Health, 2014 [accessed 2020 May 13]


Above message is taken from Centers for Disease Control (CDC). "Health Benefits of Quitting Smoking" https://www.cdc.gov/tobacco/quit_smoking/how_to_quit/benefits/#health-benefits-of-quitting-smoking. Accessed 9/1/2021.

### Benefits of Quitting Smoking

Most people know that quitting smoking is good for their physical health. Few people know that **quitting is good for their mental health** too![1]

Quitting smoking is related to a significant:[1]

**Benefit 1**

**Decrease in anxiety**

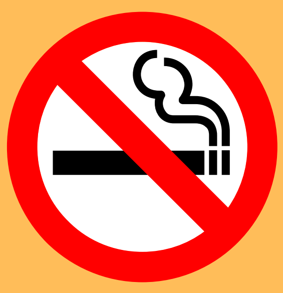

**Benefit 2**

**Decrease in depression.**

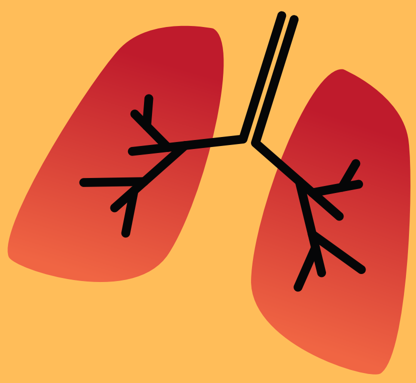

**Benefit 3**

**Decrease in stress**

**Benefit 4**

**Improvement in psychological quality of life.**

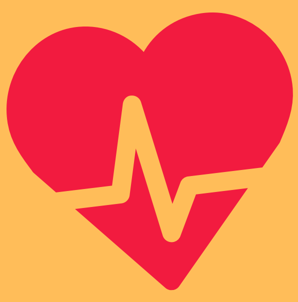

**Benefit 5**

**Increase in long-term positive mood.**

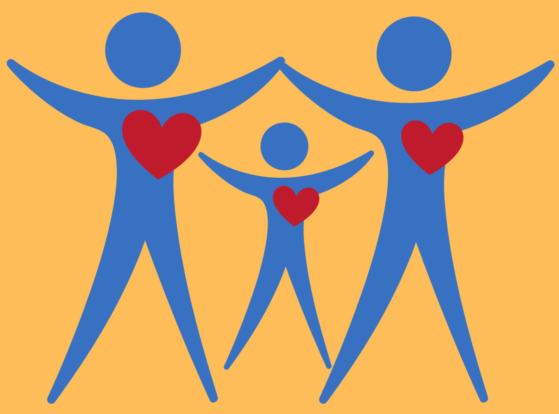

**Some people with depression or anxiety worry that their symptoms will get worse if they quit smoking. It turns out that this is not true. Research tells us that smokers have better mental health after they quit.**


1. 1. Taylor_G, McNeill_A, Girling_A, Farley_A, Lindson_N, Aveyard_P. Change in mental health after smoking cessation: systematic review and meta-analysis. BMJ 2014;348:g1151.
2. Taylor_GMJ, McNeill_A, Farley_A, Lindson_N, Aveyard_P. Smoking cessation for improving mental health. Cochrane Database of Systematic Reviews 2020, Issue 1. Art. No.: CD013522. DOI: 10.1002/14651858.CD013522.
